# Longitudinal and Geographic Trends in Perceived Racial Discrimination Among Adolescents in the U.S.: The Adolescent Brain Cognitive Development (ABCD) Study

**DOI:** 10.1101/2024.09.08.24313273

**Authors:** Christopher T. Fields, Carmen Black, Amanda J. Calhoun, Shervin Assari, Xin Zhou, Jason Nagata, Dylan G. Gee

## Abstract

This study examines longitudinal and geographic trends in perceived racial discrimination among U.S. adolescents using data from the Adolescent Brain Cognitive Development (ABCD) Study. A diverse sample of 11,868 children aged 9-10 at baseline from 22 sites across the U.S. was analyzed, assessing perceived discrimination at ages 10-11, 11-12, and 13-14 using items adapted from the Perceived Discrimination Scale. Binomial logistic regression models were used to evaluate longitudinal trends and geographic variation, adjusting for demographic factors such as race/ethnicity, parental education, and income. Results show that perceived racial discrimination increased significantly from ages 10-11 to 13-14, particularly among Black and Asian adolescents. By age 13-14, nearly half of Black adolescents and over a quarter of Asian adolescents reported discrimination. Geographic analysis revealed that Black adolescents in the Western U.S. and predominantly White affluent neighborhoods had the highest odds of perceived discrimination. Higher state-level anti-Black bias was associated with lower discrimination rates among Black adolescents but higher rates for Asian adolescents. These findings highlight the evolving nature of racial discrimination during adolescence and underscore the need for targeted interventions that address racism’s mental health impacts on adolescents, particularly in high-risk geographic and socio-economic contexts.

---

Racism is a ubiquitous force in US society, conveyed through interpersonal and structural dimensions, that impacts mental health in minoritized groups across the lifespan.^1^ A previous analysis of the Adolescent Brain Cognitive Development (ABCD) Study found that 4.8% of US children aged 10-11 years reported discrimination based on their race, ethnicity, or color.^2^ However, little is known about longitudinal trends in perceived discrimination through early adolescence or how experiences of discrimination vary across geographic regions. Such information is critical for identifying vulnerable developmental stages and high-risk communities to inform targeted interventions. Therefore, the objective of this study was to assess longitudinal and geographic variation in perceived discrimination from ages 10-11 to 13-14 years in the ABCD cohort.

## Methods

Data were drawn from the Adolescent Brain Cognitive Development (ABCD) Study (release 5.1), a diverse sample of 11,868 children aged 9-10 years at baseline, recruited from 22 sites across the United States. Perceived discrimination was assessed at the one-year (ages 10-11), two-year (ages 11-12), and four-year follow-up (ages 13-14) using items adapted from the Perceived Discrimination Scale.^3^ Respondents were categorized as perceiving discrimination if they responded affirmatively to any of 8 questions assessing unfair treatment based on race/ethnicity. Null responses were excluded from analysis.

Longitudinal trends in perceived discrimination were examined using binomial logistic regression models adjusting for parental education, household income, gender, race, and ethnicity. Geographic variation in perceived discrimination at ages 13-14 was assessed for Black and Asian adolescents, the groups with the highest rates at year 4 follow-up, using two area-level indicators: 1) a state-level index of racial animus derived from aggregated measures of explicit and implicit anti-Black bias,^4^ and 2) a census tract-level Index of Concentration at the Extremes (ICE), capturing the concentration of affluent White households relative to poor Black households.^5^ Odds ratios with Bonferroni-adjusted 95% confidence intervals (using 99.375% CIs) were estimated from models including individual and area-level factors, using propensity weights derived from the American Community Survey. Additional methodological details are provided in the eSupplement.

## Results

At ages 13-14, 45.3% of Black, 28.5% of Asian or Pacific Islander, 26.6% of Hispanic, 15.4% of Native American, and 12.6% of White adolescents reported discrimination (Table 1). In adjusted models, Black adolescents had the largest odds of perceived discrimination at 13-14 relative to White adolescents of the same age range (AOR 4.42, 99.997% CI 4.37-4.47), and Asian adolescents had the largest intra-racial increase in perceived discrimination from 10-11 to 13-14 (AOR 1.94, 99.997% CI 1.90-1.98).

**Table 1:** Longitudinal Trends in Perceived Discrimination by Race/Ethnicity from Ages 10-11 to 13-14.

|  | Year 1 | Year 2 | Year 4 | Intra-racial Year 4 /<br>Year 1 Relative AOR |
| --- | --- | --- | --- | --- |
| Total (n) | 11052 | 10759 | 4671 | - |
| White (n, %) | 996/7221<br>(13.8%) | 676/7031 (9.6%) | 412/3271<br>(12.6%) | - |
| Black (n, %) | 797/2264<br>(35.2%) | 696/2201<br>(31.6%) | 345/761 (45.3%) | - |
| Hispanic (n, %) | 515/2113<br>(24.4%) | 367/2050<br>(17.9%) | 247/927 (26.6%) | - |
| Asian or Pacific Islander (n, %) | 97/652 (14.9%) | 74/630 (11.7%) | 80/281 (28.5%) | - |
| Native American (n, %) | 67/254 (26.4%) | 34/247 (13.8%) | 18/117 (15.4%) | - |
| Other (n, %) | 162/661 (24.5%) | 134/650 (20.6%) | 71/241 (29.5%) | - |
| White (AOR) | 1 [Reference] | 1 [Reference] | 1 [Reference] | 0.82 (0.81-0.83) |
| Black (AOR) | 2.31 (2.29-2.32) | 2.85 (2.83-2.87) | 4.42 (4.37-4.47) | 1.45 (1.43-1.46) |
| Hispanic (AOR) | 1.28 (1.26-1.28) | 1.18 (1.17-1.19) | 1.55 (1.53-1.57) | 1.09 (1.08-1.10) |
| Asian or Pacific Islander (AOR) | 1.49 (1.47-1.51) | 1.48 (1.46-1.51) | 2.88 (2.83-2.93) | 1.94 (1.90-1.98) |
| Native American (AOR) | 1.68 (1.66-1.71) | 1.16 (1.13-1.18) | 0.92 (0.89-0.94) | 0.42 (0.41-0.44) |
| Other (AOR) | 1.38 (1.36-1.39) | 1.71 (1.69-1.73) | 1.78 (1.75-1.81) | 1.23 (1.21-1.25) |

Adjusted odds ratios (AOR) for perceived discrimination for Years 1, 2, or 4 are relative to White participants
from the same year, using factors race (White), gender (non-Male), ethnicity (non-Hispanic), combined parental income (>\$75K), and parental education (any guardian having four-year college degree), adjusted using American Community Survey (ACS) weights. Intra-racial Year 4 to Year 1 Relative AOR is calculated within racial/ethnic group, comparing Year 4 to Year 1 (e.g., Black AOR Year 4 vs Black AOR Year 1). Null responders to all racial/ethnic discrimination questions were excluded. Unweighted number of participants reported as participants affirming discrimination versus total number of participants attributed to the referenced racial/ethnic group. AORs are reported with Bonferroni-corrected 95% CI (using 99.997% CIs).

At ages 13-14, Black adolescents in the West had the highest adjusted odds of perceived discrimination compared to all other regional by racial/ethnic groups (AOR 7.42, 99.997% CI 7.16-7.70) (Table 2). Likewise, Black adolescents living in high ICE census tracts with greater concentrations of affluent White households had higher odds of perceived discrimination in comparison to any other combination of ICE levels and racial/ethnic groupings (AOR 12.71, 99.997% CI 12.29-13.16). Higher state-level anti-Black bias was associated with lower rates of perceived discrimination for Black adolescents, but with higher rates of perceived discrimination for Asian adolescents.

**Table 2:** Geographic Variation in Perceived Discrimination Among Black and Asian Adolescents at Ages 13-14.

|  | Black | Asian |
| --- | --- | --- |
| Northeast (n, %) | 72/151 (47.7%) | 7/23 (30.4%) |
| Midwest (n, %) | 75/181 (41.4%) | 14/40 (35.0%) |
| South (n, %) | 148/339 (43.7%) | 14/47 (29.8%) |
| West (n, %) | 50/90 (55.6%) | 45/171 (26.3%) |
| High ICE Disparity / Wealthy White (n, %) | 46/83 (55.4%) | 39/134 (29.1%) |
| High ICE Disparity / Poor Black (n, %) | 43/96 (44.8%) | no participants |
| Low ICE Disparity / Wealthy White (n, %) | 130/294 (44.2%) | 34/129 (26.4%) |
| Low ICE Disparity / Poor Black (n, %) | 119/260 (45.8%) | 4/10 (40%) |
| High State-Level Anti-Black Bias (n. %) | 192/436 (44.0%) | 20/60 (33.3%) |
| Low State-Level Anti-Black Bias (n. %) | 153/325 (47.1%) | 60/221 (27.1%) |
| Northeast (AOR) | 4.94 (4.82-5.06) | 3.15 (2.99-3.32) |
| Midwest (AOR) | 3.87 (3.78-3.96) | 4.37 (4.22-4.53) |
| South (AOR) | 4.15 (4.08-4.22) | 2.72 (2.63-2.82) |
| West (AOR) | 7.42 (7.16-7.70) | 2.55 (2.48-2.61) |
| High ICE Disparity / Wealthy White (AOR) | 12.71 (12.29-13.16) | 4.82 (4.69-4.94) |
| High ICE Disparity / Poor Black (AOR) | 7.30 (7.11-7.50) | - |
| Low ICE Disparity / Wealthy White (AOR) | 6.43 (6.31-6.55) | 3.78 (3.69-3.88) |
| Low ICE Disparity / Poor Black (AOR) | 6.54 (6.41-6.67) | 5.50 (5.14-5.88) |
| High State-Level Anti-Black Bias (AOR) | 4.85 (4.78-4.93) | 4.50 (4.37-4.64) |
| Low State-Level Anti-Black Bias (AOR) | 5.51 (5.42-5.61) | 2.98 (2.92-3.04) |

Adjusted odds ratios (AOR) for perceived discrimination among Black and Asian adolescents at ages 13-14 by geographic region, neighborhood socioeconomic composition (Index of Concentration at the Extremes; ICE), and state-level anti-Black bias. AORs are adjusted for individual-level factors including gender, parental income, and parental education, adjusted using ACS weights. Unweighted number of participants reported as participants affirming discrimination versus total number of participants. AORs are reported with Bonferroni-adjusted 95% confidence intervals.

## Discussion

In this large, diverse sample, perceived discrimination increased from ages 10-11 to 13-14, particularly among Black and Asian adolescents. By ages 13-14, nearly half of Black and over a quarter of Asian adolescents reported discrimination, with Black adolescents reporting the highest rates in the West and in advantaged, predominantly White neighborhoods. These findings align with prior research suggesting that higher income is associated with more perceived discrimination among Black adolescents in predominantly White areas.^6^ The pervasiveness of racism-related experiences among adolescents underscores the need for early prevention strategies in clinical and community settings.

## Supporting information

eSupplement

## Data Availability

ABCD data are available to qualified researchers through the NIMH Data Archive (NDA). The data used in this report are from the fast track data release 5.1. The raw data can be accessed at https://nda.nih.gov/study.html?id=2313. The data dictionary for the ABCD study is available at https://data-dict.abcdstudy.org/, and additional information on the measures assessed in the ABCD study can be found at https://wiki.abcdstudy.org/release-notes/start-page.html. Instructions on how to obtain NDA Data Use Certification are provided at https://nda.nih.gov/nda/access-data-info.
Statistical and analytical code will be made available to researchers with an approved NDA Data Use Certificate upon publication. Requests for access to the code should be directed to. The ABCD data are available for analyses approved by NDA, and all access requires a Data Use Certificate (DUC) issued by NDA.

https://data-dict.abcdstudy.org/

https://wiki.abcdstudy.org/release-notes/start-page.html

https://nda.nih.gov/nda/access-data-info

## Author Contributions

Dr Fields had full access to all of the data in the study and takes responsibility for the integrity of the data and the accuracy of the data analysis.

*Concept and design:* All authors.

*Acquisition, analysis, or interpretation of data:* Fields, Nagata, Gee.

*Drafting of the manuscript:* Fields, Black, Calhoun, Assari.

*Statistical analysis:* Fields, Nagata, Zhou, Gee. *Administrative, technical, or material support:* Nagata, Gee. *Supervision:* Black, Assari, Gee.

## Additional Information

A complete listing of the ABCD study investigators can be found at https://abcdstudy.org/principal-investigators.html.

