## Supplementary material for "Longitudinal and Geographic Trends in Perceived Racial Discrimination Among Adolescents in the U.S.: The Adolescent Brain Cognitive Development (ABCD) Study": eSupplement

Supplementary Methods

All analyses were completed using the 5.1 release of the Adolescent Brain Cognitive Development (ABCD) study, which encompasses a total of 11,868 participants recruited from 22 sites across the United States. These sites were geographically diverse, with 5 sites located in the Northeast region, 4 in the Midwest, 6 in the South, and 7 in the West. The ABCD study began its recruitment process in September 2016, initially focusing on children between the ages of 9 and 10 years old. Prior to their participation, the children provided their assent, and their parents or legal guardians gave informed consent. All study procedures were reviewed and approved by either the respective local Institutional Review Boards or by a centralized Institutional Review Board at the University of California, San Diego.

The ABCD consortium collected demographic information from the baseline assessment. However, as the questions pertaining to perceived discrimination were not included in the baseline or the three-year follow-up assessment, we gathered these data from the one-year, two-year, and four-year follow-up assessments.

Data were preprocessed with Tableau Prep Builder (v.2024 1.0, Salesforce, San Francisco, CA) and statistical analyses were completed with R (v4.3.2).

Given the zero-inflated nature of the data, our primary outcome measure (“perceived discrimination”) is the combined binary outcome of perceived discrimination (“Yes” or “Sometimes”/”Often”/”Very Often”) across any of these eight questions:

1) Answered “yes”: “In the past 12 months, have you felt discriminated against: because of your race, ethnicity, or color? Definition of ethnicity: groups of people who have the same customs, or origin”
2) Answered “3” (“sometimes”), “4” (“Often”), or “5” (“Very often”): “How often do the following people treat you unfairly or negatively because of your ethnic background? Teachers”
3) Answered “3” (“sometimes”), “4” (“Often”), or “5” (“Very often”): “How often do the following people treat you unfairly or negatively because of your ethnic background? Other adults outside school”
4) Answered “3” (“sometimes”), “4” (“Often”), or “5” (“Very often”): “How often do the following people treat you unfairly or negatively because of your ethnic background? Other students”
5) Answered “3” (“sometimes”), “4” (“Often”), or “5” (“Very often”): “I feel that others behave in an unfair or negative way toward my ethnic group”
6) Answered “3” (“sometimes”), “4” (“Often”), or “5” (“Very often”): “I feel that I am not wanted in American society”
7) Answered “3” (“sometimes”), “4” (“Often”), or “5” (“Very often”): “I don't feel accepted by other Americans”
8) Answered “3” (“sometimes”), “4” (“Often”), or “5” (“Very often”): “I feel that other Americans have something against me”

Subjects who are consistently marked as “null”, “don’t know”, or “refuse to answer” across all eight questions were designated as null participants and were excluded from analysis. If a subject answered “Yes” to Question 1 or “3”, “4”, or “5” to Questions 2-8, they were marked as “Yes” for “Perceived Discrimination.” (Responses of “No” to Question 1 or “1” [“Almost Never”] or “2” [“Rarely”] to Questions 2-8 were designated as “No” for “Perceived Discrimination.”)

No data were missing regarding regional assignment. ‘77’ (“Refuse to Answer”) and ‘99’ (“Don’t Know”) and null responses for race were excluded. Missing values for parental education, parental income, and gender were coded under “No college degree or education not reported”, “<$75K or parental income not reported”, and “non-male or gender not reported” respectively. For models 5-8 (see below), null values for ICE Factors and the State Racism Factors were excluded from analyses.

For geographic trends, we included two contextual factors: a state-level racism factor developed by Hatzenbuehler et al. and a census tract-level Index of Concentration at the Extremes for race/ethnicity and income (ICE-R) developed by Krieger et al. The state racism factor comprised 31 items assessing aggregated attitudes related to race and racial prejudice, which were obtained from three sources: Project Implicit (pooled across years 2002–2017), the General Social Survey (pooled across years 1973–2014), and the American National Election Survey (pooled across years 1992–2016). These items collectively assessed different components of racial prejudice, including general attitudes toward Black people, the perceived impact of discrimination on the lives of Black people, the existence of racial prejudice, and endorsement of racial stereotypes. State racism scores below 0 were categorized as “low anti-black bias” and scores 0 and above were categorized as “high anti-black bias”. The ICE-R measures the extent to which the residents in a census tract are concentrated into the extremes of privilege and deprivation based on race/ethnicity and income. Specifically, we used the ICE-BW variant which is calculated as the concentration of affluent (top 20% of household income) non-Hispanic white households compared to the concentration of poor (bottom 20% of household income) non-Hispanic Black households. Including both the state racism factor and the census tract ICE-BW allowed us to examine the impact of structural racism and racialized economic segregation at the state and local neighborhood levels on the outcomes of interest. ICE-BW scores with an absolute value of 0.3 and above were categorized as “high disparity” areas and below 0.3 were categorized as “low disparity” areas. ICE-BW scores 0 and above were categorized as “wealthy white biased” areas, whereas ICE-BW scores below 0 were categorized as “poor black biased” areas.

Adjusted odds ratios (AORs) were calculated from eight binomial logistic regression models, all built with maximum likelihood estimation. All models were built with intercepts, using the general equation: $\log\left( \frac{p}{1-p} \right)=\beta_{0}+\beta_{1}X_{1}+\beta_{2}X_{2}+\ldots+\beta_{n}X_{n}$, where p is the probability of perceived discrimination, $\beta_{0}$ is the intercept, $(\beta_{1},\ldots,\beta_{n})$ are the coefficients for independent variables ($X_{1},\ldots, X_{n})$ variables.

Bonferroni correction was done across fifteen models. (Results for Hispanic ethnicity and White, Native-American, and Other races not reported in Table 2.) The alpha level was set to = 0.05/15 = 0.00333. 95% confidence intervals for AORs were corrected to 99.997% confidence intervals.

Models were built with a combination of the following factors (levels; reference):
Parental Four-Year College Degree (Four Year College Degree, No Four Year College Degree; reference = Four Year College Degree)

Parental Income (>$75K, <$75K; reference = >$75K)

Gender (Non-Male, Male; reference = Male)

Race (White, Black, Asian or Pacific Islander, Native American, Other; reference = White)

Ethnicity (Non-Hispanic, Hispanic; reference = Non-Hispanic)

Region (West, Northeast, Midwest, South; reference = West)

ICE-BW (High Disparity Wealthy White Biased, High Disparity Poor Black Biased, Low Disparity Wealthy White Biased, Low Disparity Poor Black Biased; reference = High Disparity Wealthy White Biased)

State Racism Factor (Low Anti-Black Bias, High Anti-Black Bias; reference = Low Anti-Black Bias)

Site (site01 … site22; reference = site01)

The models reported for table 1:

Model 1 (Year 1 Perceived Discrimination): ParentalEducation, ParentalIncome, Gender, Ethnicity, Race, Site

Model 2 (Year 2 Perceived Discrimination): ParentalEducation, ParentalIncome, Gender, Ethnicity, Race, Site

Model 3 (Year 4 Perceived Discrimination): ParentalEducation, ParentalIncome, Gender, Ethnicity, Race, Site

Model 4 (White longitudinal model): ParentalEducation, ParentalIncome, Gender, Ethnicity, Year, Site

Model 5 (Black longitudinal model): ParentalEducation, ParentalIncome, Gender, Ethnicity, Year, Site

Model 6 (Asian or Pacific Islander longitudinal model): ParentalEducation, ParentalIncome, Gender, Ethnicity, Year, Site

Model 7 (Native American longitudinal model): ParentalEducation, ParentalIncome, Gender, Ethnicity, Year, Site

Model 8 (Other longitudinal model): ParentalEducation, ParentalIncome, Gender, Ethnicity, Year, Site

Model 9 (Hispanic longitudinal model): ParentalEducation, ParentalIncome, Gender, Ethnicity, Year, Site

Model 2 Factors: ParentalEducation, ParentalIncome, Gender, Race, EthnicityYear

For table 2 (restricted to Year 4 data):

Model 10 Factors: ParentalEducation, ParentalIncome, Gender, Race * Region

Model 11 Factors: ParentalEducation, ParentalIncome, Gender, Ethnicity * Region

Model 12 Factors: ParentalEducation, ParentalIncome, Gender, Race * ICE

Model 13 Factors: ParentalEducation, ParentalIncome, Gender, Ethnicity * ICE

Model 14 Factors: ParentalEducation, ParentalIncome, Gender, Ethnicity, Race * StateRacismFactor

Model 15 Factors: ParentalEducation, ParentalIncome, Gender, Ethnicity * StateRacismFactor

Supplementary Table: Site code for location and region

| SiteNumber | Abbreviation | Full Site Name | City and State | Region |
| --- | --- | --- | --- | --- |
| site01 | CHLA | Children's Hospital Los Angeles | Los Angeles, CA | West |
| site02 | CUB | University of Colorado Boulder | Boulder, CO | West |
| site03 | FIU | Florida International University | Miami, FL | South |
| site04 | LIBR | Laureate Institute for Brain Research | Tulsa, OK | South |
| site05 | MUSC | Medical University of South Carolina | Columbia, SC | South |
| site06 | OHSU | Oregon Health & Science University | Portland, OR | West |
| site07 | ROC | University of Rochester | Rochester, NY | Northeast |
| site08 | SRI | SRI International | Menlo Park, CA | West |
| site09 | UCLA | UCLA | Los Angeles, CA | West |
| site10 | UCSD | University of California San Diego | San Diego, CA | West |
| site11 | UFL | University of Florida | Gainsville, FL | South |
| site12 | UMB | University of Maryland at Baltimore School of Medicine | Baltimore, MD | South |
| site13 | UMICH | University of Michigan | Ann Arbor, MI | Midwest |
| site14 | UMN | University of Minnesota | Minneapolis, MN | Midwest |
| site15 | UPMC | University of Pittsburgh Medical Center | Pittsburgh, Pa | Northeast |
| site16 | UTAH | University of Utah | Salt Lake City, UT | West |
| site17 | UVM | University of Vermont | Burlington, VT | Northeast |
| site18 | UWM | University of Wisconsin-Milwaukee | Milwaukee, WI | Midwest |
| site19 | VCU | Virginia Commonwealth University | Richmond, VA | South |
| site20 | WUSTL | Washington University in St. Louis | St. Louis, MO | Midwest |
| site21 | YALE | Yale University | New Haven, CT | Northeast |
| site22 | MSSM | Mt. Sinai School of Medicine | New York, NY | Northeast |
